# A Probabilistic Synthesis of Malaria Epidemiology: Exposure, Infection, Parasite Densities, and Detection

**DOI:** 10.1101/2025.03.24.25324561

**Authors:** John M. Henry, Austin R. Carter, Sean L. Wu, David L. Smith

## Abstract

The epidemiology of Plasmodium falciparum malaria presents a unique set of challenges due to the complicated dynamics of infection, immunity, disease, and detection. Studies of malaria epidemiology commonly measure malaria parasite densities or prevalence, but since malaria is so complex with so many factors to consider, a complete mathematical synthesis of malaria epidemiology has been elusive. Here, we take a new approach. From a simple model of malaria exposure and infection in human cohorts as they age, we develop random variables describing the multiplicity of infection (MoI) and the age of infection (AoI). Next, using the MoI and AoI distributions, we develop random variables describing parasite densities, parasite counts, and detection. We also derived a random variable describing the age of the youngest infection (AoY), which can be used to compute approximate parasite densities in complex infections. Finally, we derive a simple system of differential equations with hybrid variables that track the mean MoI, AoI and AoY, and we show it matches the complex probabilistic system with reasonable accuracy. We can thus compute the state of any individual chosen at random from the population in two ways. The same approach – pairing random variables and hybrid models – can be extended to model other features of malaria epidemiology, including disease, malaria immunity, treatment and chemoprotection, and infectiousness. The computational simplicity of hybrid models has some advantages over compartmental models and stochastic individual-based models, and with the supporting probabilistic framework, provide a sound basis for a synthesis of observational malaria epidemiology.

**Significance:** The probabilistic framework presented here represents a step forward in the theory for observational malaria epidemiology. Using these methods, one can connect the heterogeneous dynamics of individual infections to both observational and process uncertainty in estimates of population-level metrics such as cross-sectional surveys of prevalence by light microscopy. This serves as a basis for more complex simulation studies using the present models and extensions that include factors such as fever, treatment, and transmission.

---

The epidemiology of human malaria caused by infection with *Plasmodium falciparum* is complex. Exposure to *P. falciparum*, as measured by the annual entomological inoculation rate (aEIR), varies by several orders of magnitude seasonally, across years, and among locations (1–3). The duration of detectable parasitemia for simple malaria infections varies from a few weeks to more than a year, and parasite densities are highly variable among individuals in populations depending on their age, location, and the time of year (4–6). In malaria therapy – a peculiar set of observations in which exposure was fully known – detection, disease and infectiousness all varied with the age of infection (AoI) (7, 8). While these studies provide a rich source of data describing malaria, controlled infections in those institutionalized patient populations differed from natural settings in many ways. In natural settings, neither immunity nor infection prevents reinfection (also called superinfection), so malaria infections can be complex – the number of distinct parasite lineages has been called the multiplicity of infection (MoI) (9–11), and a person could experience dozens of bouts of moderate to severe malaria over a lifetime (3, 12). Acquired and innate immunity modify infection and the risk and severity of disease, but protection is weak, has a poor memory, and accumulates slowly with age and exposure (13–15). The consensus is that malaria immunity varies in some way with age, cumulative exposure, and recent exposure (3, 12, 16), but there are some important data gaps and lingering uncertainty. A synthesis is needed.

An important feature of malaria is the long, variable time course of infection (5, 6, 17). After the infective bite and a short period in the liver, parasites begin a cycle of asexual replication in red blood cells, all while their numbers are increasing geometrically (18). Immunity to malaria and other factors stop the geometric growth phase, and then the infection begins a long and variable chronic phase, during which parasite densities tend to decline and are detected less frequently until eventually the infection ends (8). Untreated malaria infections can, nevertheless, persist for a few weeks to several years. The time course of malaria infections is highly variable over the time course of an infection, and the features of those infections vary over a lifetime. Substantial attention has focused on the *var* genes, and *var* gene switching as a mechanism that allows parasite infections to persist, but parasites can also modify gene transcription in other ways (17, 19), and there are other genes that play an important role in development of immunity. The net effect of the interplay between parasites and the human immune response is that parasite densities are highly variable over the time course of an infection, and predictable only in a statistical sense (8, 17). Throughout an infection, the probability of detecting parasites varies, so accurate measurement of the distribution of basic features of malaria, such as the duration of infections, has been difficult. Parasite densities in blood fluctuate over several orders of magnitude, and only a fraction of blood is examined. The sensitivity of parasite counts or other estimates of parasite density and malaria prevalence varies across diagnostic methods (20, 21). Available data thus measure the duration of patentcy, but evidence for very long carriage times comes from infections that have recrudesced after persisting for decades.

A second feature of malaria epidemiology is re-exposure and superinfection. Acquired immunity to malaria does not prevent reinfection, so after subsequent exposure, so multiple, distinct parasite lineages can coinfect a single host. It is also possible for a single infective bite to transmit multiple distinct parasite clones. Several studies have attempted to measure the MoI, but the metric is difficult to interpret given the probability of detection (9). Understanding parasite density distributions in complex infections (*i*.*e*., those with MoI*>* 1) presents a daunting challenge, in part, because parasite densities are so variable. With highly variable infections, highdensity infections in one sample can mask other low-density infections. The complexity of infections may be partially revealed by sampling the same person repeatedly over time (22, 23).

The drive to understand malaria has led to a proliferation of models that address various aspects of malaria, including simple models exploring various aspects of transmission, and comprehensive and synthetic studies using individual-based and intra-host simulation models of malaria infections (24–27). Because the number of factors affecting malaria is large, the state space describing malaria is high-dimensional. Given the uncertainty about malaria epidemiology and its complexity, a conceptual and quantitative synthesis of malaria epidemiology must be developed around some theory and mathematical models. A practical challenge, from a mathematical or computational perspective, has been how to represent the multi-dimensional state space for infection and immunity – as models consider more factors, the size of the state space expands combinatorically and becomes difficult to manage. One way of handling the complex state space has been through development of individual-based simulation models, which replicate the basic patterns in models that simultaneously deal with the complex time course of infections, superinfection, and waxing and waning immunity (28–36). While IBMs are a useful computational tool for many purposes, they are often presented as black boxes due to the complexity of their design and the probabilistic nature of their output. What is needed is a study of relevant detail to help identify the critical dimensions without losing information and prioritize studies that could fill critical gaps (**?**).

Here, we take a probabilistic approach to understanding malaria epidemiology, including parasite density distributions, by formulating random variables that track the AoI, the MoI, and parasite density distributions in human birth cohorts as they age. In the simple infections observed in malaria therapy patients, most aspects of infections were well described statistically by the AoI, the time elapsed since the infective bite for each parasite clone infecting a host: the fraction persisting, the fraction still detectable, the mean and variance of logged parasite densities, the risk of fever and disease, and infectiousness (8). Using the AoI a unifying concept, we developed models of understand the density distributions of parasite clones in cohorts of humans as they age; these distribution link exposure, the MoI, the AoI, parasite densities in complex infections, and detection. Using this approach, we show that malaria epidemiology, including parasite densities, can be understood in terms of a new random variable describing the age of the youngest infection (AoY). This approach does not rely on stochastic simulations, and we have found it more amenable to analysis. Finally, for analysis and synthesis, we developed hybrid variables describing the mean AoI and mean AoY, and we show how a simple systems of differential equations holds most of the information about malaria infection dynamics, so computationally simple models of malaria epidemiology can be used to understand complex patterns minimal information loss. Altogether, the methods present a basis for a conceptual synthesis of observational malaria epidemiology.

## Dynamical Models for Cohorts

In the following, we analyze a simple model for malaria infection in a host cohort as it ages. The analysis extends a queuing model for super-infection called *M/M/*∞ (Fig 1), which has played an important role in malaria epidemiology (37–40). A core assumption of the model is that infections clear independently, and in the spirit of that model, we also assume that parasite densities over the time courses of two or more parasite infections are also independent. In this study, we have focused on understanding how malaria epidemiology changes in young children as they age, depending on exposure.

**Fig. 1.**
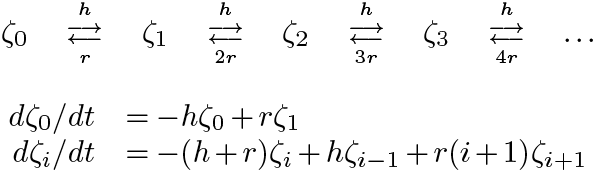
A diagram of the queuing model *M/M/*∞ (top), and the master equations (bottom), in which *ζ*_*i*_ denotes the proportion of the population with MoI = *i*. MoI increments by one if an infection occurs at rate *h* and decrements by one if an infection clears at rate *ri*.

In the following, we let *a* denote the age of a host birth cohort as it ages over time. A *parasite clone* is defined to include all the progeny of the parasites arising from a single infective mosquito bite. We are thus assuming that each bite would transmit a single clone, an assumption that could be relaxed in subsequent analyses. We let *α* denote the AoI: the age of a parasite clone, or the time elapsed since the infective bite that initiated the clonal infection.

The random variables are all computed conditioned on exposure, which we define as the force of infection (FoI), the hazard rate for infection experienced by a cohort of susceptible humans as it ages. Starting from a model for the infection dynamics, we developed random variables describing the MoI, the AoI, and parasite densities as a function of the FoI (see Fig 2). We then model the observational process, which translates parasite densities into parasite counts and measures of prevalence. A synthesis was achieved through two extensions: first, we defined a new random variable describing the age of the youngest infection (AoY); next, we derived hybrid variables describing the mean AoI and the mean AoY.

**Fig. 2.**
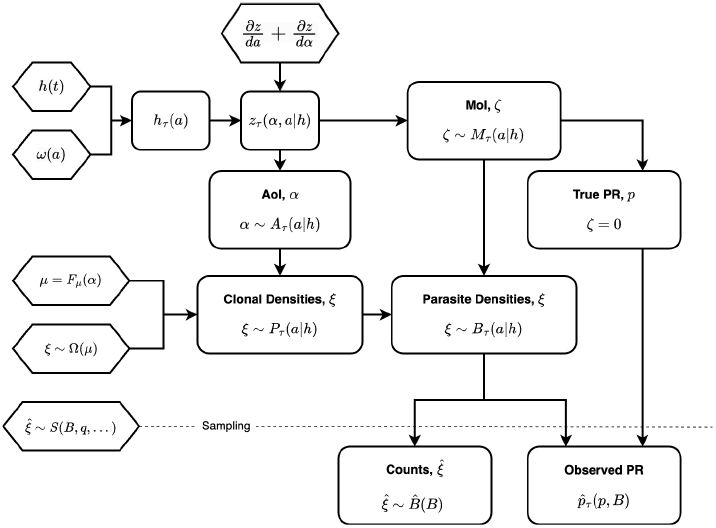
An overview of the probabilistic model and development of the random variables. Model assumptions are drawn as hexagons. The random variables are developed around a model for exposure and infection dynamics. We pass the daily FoI in the population, *h*(*t*), and an assumption about exposure by age, *ω*(*a*), so that the model input is the daily FoI for a particular cohort as it ages, *h*_*τ*_ (*a*). We then solve the equation describing the density of parasite clones of age *α*, distributed among a cohort of hosts as it ages, *z*(*α, a*|*h*). We compute random variables describing the MoI, *M*_*τ*_ (*a* | *h*), and the AoI *A*_*τ*_ (*a* | *h*). We assume mean log_10_ parasite densities, *ξ*, are a function of the AoI (*F*_*µ*_(*α*)) with a probability density function given by Ω(*µ*). Assuming independence, we then derive random variables describing the clonal density of parasites in blood, *P*_*τ*_ (*a*|*h*), and total parasite densities, summed over all clonal infections, *B*_*τ*_ (*a*| *h*). Under a model for sampling parasites from a fraction *q* of the blood, *S*(*B, q*,…), We compute the observed PR, 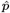,and the distribution of counts, 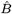

The primary computational methods for the paper are found in open source R package on github, called `ramp.falciparum.` Derivations for most of the formulas and some proofs are found in the Supporting Information. Vignettes in `ramp.falciparum` walk through the derivations, provide pseudo-code to compute most random variables, and provide numerical validation for most of the claims (https://github.com/dd-harp/ramp.falciparum). The software was designed to be both modular and extensible, and there are plans to extend this probabilistic approach to model immunity, disease, treatment and chemoprotection, and other factors.

### Exposure and Infection

Differences in the rate of exposure by age and season are modelled through the FoI, a time- and age-dependent average hazard rate for new infections in a susceptible population. To make the analysis as general and applicable as possible, we have developed methods to generate a broad class of functions to describe the FoI in a population over time, *h*(*t*). In doing so, we are explicitly reducing the FoI to a model input, or a *trace function*. In many studies with mechanistic models, the FoI computed as the result of a dynamical process reflecting parasite transmission from humans to mosquitoes, and blood feeding by mosquitoes. These trace-driven simulations are one way to decouple different parts of a simulation when one aspect of a simulation can be replaced by an external input (trace) from other parts of the simulation on which it depends (41**?**). Using this approach, we can investigate malaria under almost any pattern of exposure without the need to specify exactly how that pattern of exposure might have arisen.

### The Force of Infection

The GitHub R package, `ramp.falciparum`, includes a flexible yet “realistic” set of functions to model exposure (Fig 3 and Supporting Information). We let 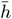 be a parameter that sets the overall level of exposure. We assume exposure has a seasonal pattern, *S*(*t*), and a temporal trend, *T* (*t*):

**Fig. 3.**
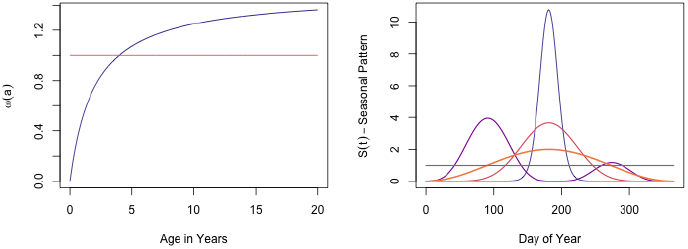
FoI trace functions developed to simulate exposure combine four elements: a parameter that sets the overall intensity,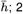) functions that modify exposure by age, *ω*(*a*); 3) functions that describe the seasonal pattern, *S*(*t*); and 4) and a long-term trend, *T* (*t*). The left panel illustrates representative functions for agemodification, and the right panel shows representative seasonal patterns.

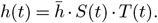

Motivated by the evidence for age-dependent exposure, the rate of exposure in the cohort is assumed to change with age. We use the age-weighting function, *ω*(*a*), consistent with the available evidence (42, 43). Each cohort is indexed by its birth day, *τ*. The FoI at age *a* for the cohort born on day *τ*, which is at time *t* = *a* + *τ*, is:

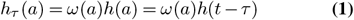

We can thus develop arbitrary exposure patterns and compare outcomes for cohorts of different ages in the same population, of the same age at different times.

### Infection Dynamics

Let *z*(*α, a*) denote the density of parasite clonal infections of age *α* in a host cohort of age *a*. We assume infections clear at the constant rate *r*. Since infections in *M/M/*∞ are independent, we can track the dynamics for the AoI of all parasite clones with the equation,

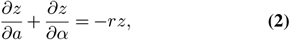

with the boundary condition *z*_*τ*_ (0, *a*) = *h*_*τ*_ (*a*). We note that the age of the host birth cohort sets an upper limit for the age of the parasite clones, so 0 *< α < a*. The solution to Eq. 2, which describes density of infections of age *α* in a host cohort of age *a*, is given by the formula:

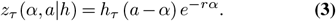

In the Supporting Information, we prove that Eq. 3 is a solution to Eq. 2.

### Age and Multiplicity of Infection

Under the assumptions of *M/M/*∞, all the parasite clones are distributed independently among hosts. Importantly, the mean MoI in the cohort at age *a* is the integral of this density function:

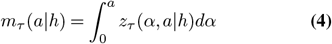

Elsewhere, it has been shown that the distribution of the MoI converges to a Poisson (44). Since the MoI is zero at birth (a degenerate Poisson distribution), the distribution of the MoI remains Poisson as the cohort ages (in this model), and mean MoI, *m*_*τ*_ (*a*) completely describes the probability density function for the MoI:

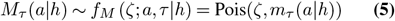

We let *F*_*M*_ denote the CDF, and 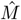 the random number function. In `ramp.falciparum,` we verify that, for the same function *h*_*τ*_ (*a*), that Eq. 4 has the same mean as *M/M/*∞.

Since *f*_*M*_ is Poisson, true prevalence (*p*) is the proportion with MoI greater than 0, which can be computed directly from *m*_*τ*_ (*a*):

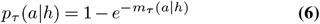

While *z*_*τ*_ (*α, a*) describes the density of clonal infections in the population, a random variable describing the density of the AoI, *A*, within hosts in particular cohort at a given age *a* is:

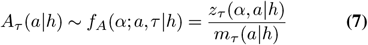

We let *F*_*A*_ denote the CDF, and *Â* the random number function. From a trace function, *h*_*τ*_ (*a*), we can directly compute and visualize the full probability distribution function for the AoI and the MoI in a cohort as it ages (Fig 4). In `ramp.falciparum`, we developed functions to compute the probability density functions, cumulative distribution functions, and random numbers for a cohort of age *a* born on day *τ* directly from *h*(*t*).

**Fig. 4.**
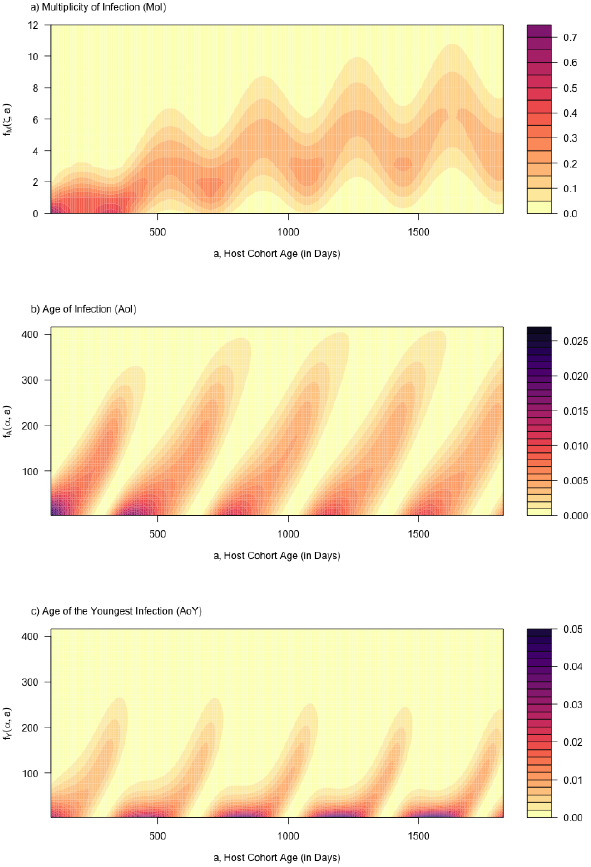
The distribution of the a) multiplicity of infection (MoI), b) the age of infection (AoI), and c) age of the youngest clonal infection (AoY) in a cohort as it ages from birth up through 5 years old. The FoI was sinusoidal, with a population mean aFoI of 5 infections, per year.

An important feature of this model is that the distribution of the AoI is affected by the seasonal pattern and age, but not by its overall intensity. Later, we will show that the patterns tend to be driven by youngest infections, which is affected by overall intensity (Fig 4b,c).

### Parasite Densities in Infections

Parasite densities in the blood are often measured clinically and reported as parasite counts, and counts or infection prevalence are commonly used as covariates in observational studies. Early studies of malaria parasite counts observed enormous variability in a single host over time as well as among hosts (4, 5). Evidence describing parasite densities improved in quality, granularity, and quantity during malaria therapy, when patients with neuro-syphilis were deliberately infected with malaria and carefully monitored (45, 46). More recently, a vast amount of detail about the early stages of infection has been learned through controlled human malaria infections (CHMI), where patients are deliberately exposed, often as part of clinical trials for developmental phase drugs or vaccines (18, 47). Here, we present a model for parasite densities and counts using the concept of the AoI, and we fit it to the malaria therapy data using maximum likelihood estimation.

In the malaria therapy data, the variance in parasite counts was a power-function of the mean, suggesting that the variance in logged parasite would be a simple linear function of the logged mean. We thus developed a realistic probability distribution function that explicitly modeled the variance as a function of the mean. Under a model of detection, we fitted the model to the malariotherapy data (Fig. 5).

**Fig. 5.**
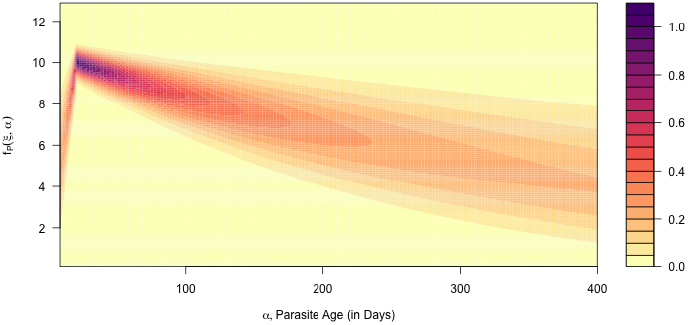
The surface plots the model fit to the malaria-therapy data (Supplement-S5).

To model parasite density distributions, we proceed in several steps: first, we formulate models for mean log_10_ parasite densities in a parasite clonal infection (denoted *µ*) as a function of its AoI (*α*) or *F*_*µ*_(*α*); second, we develop a family of probability distributions describing the number of infected red blood cells (log iRBCs, *ξ*) infected by each clone (denoted Ω) as a function of the mean (*µ*), or *ξ* ~ Ω(*µ*); third, we combine these two models, and we develop a random variable describing the density of parasite clones infecting a host cohort of age *a* born on day *τ*, denoted *P*_*τ*_ (*a* | *h*) (Fig 5); and fourth, we develop a new random variable that sums parasite densities over the MoI to describe total iRBCs in an infection, *B*_*τ*_ (*a*|*h*) (Fig. 6).

**Fig. 6.**
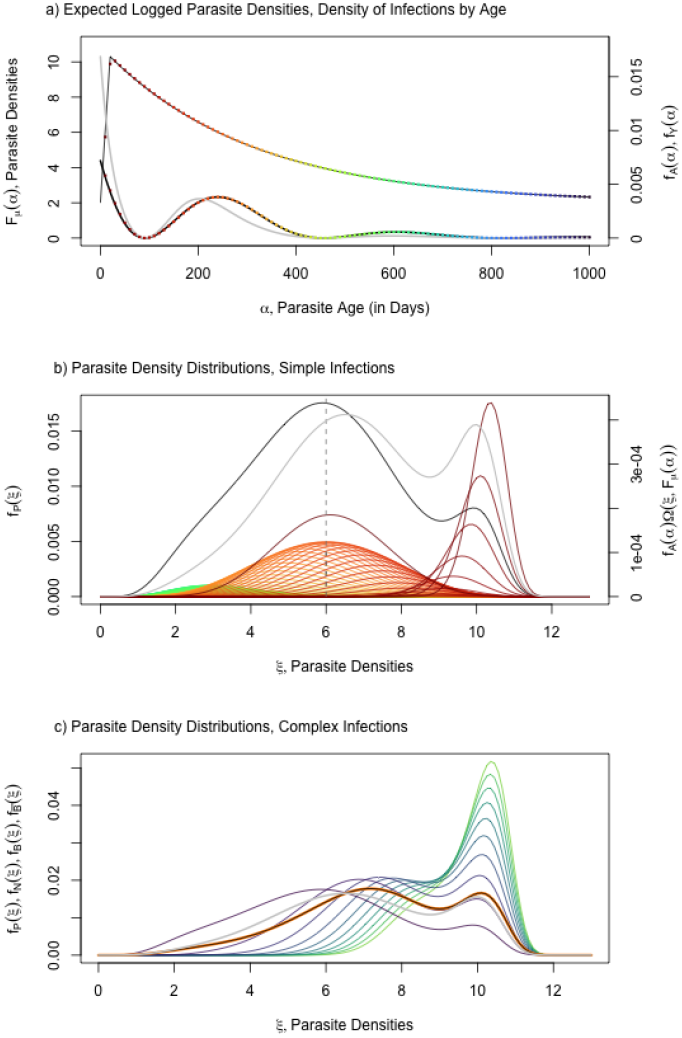
Top) the expected value for parasite densities, *F*_*µ*_(*α*) (black, Eq. 8) and the distribution of AoI, *f*_*A*_(*α*) (black line with colored points, Eq. 7), and the AoY, *f*_*Y*_ (*α*) (grey line) for one particular cohort at 5 years old. Bottom) The total distribution of parasite clonal densities across all parasite infections, *f*_*P*_ (*α*), integrates parasite density distributions, Ω(*µ*) with expected value *µ* = *F*_*µ*_(*α*) (top panel), over the distribution of the AoI. For each colored point in the top, we plot the density distribution of of parasites clones, Ω, of each age from a host cohort. c) Parasite density distribution for a clone (*f*_*P*_) for MoI= *N* (*f*_*N*_, N=2,…,10, colored purple to green), for the full distribution (*f*_*B*_, orange). We also plot 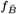 (grey).

### Time Course of Infections

Consistent with most of observational epidemiology, and based on an analysis of malariotherapy data (8), we assume that the densities of each parasite clone vary probabilistically with its age, *α*. Generically, we let *µ* = *F*_*µ*_(*α*) denote some function describing the expected mean log_10_ parasite clonal densities in a simple infection as a function of the AoI. The functions could be modified to include variables describing cumulative exposure, recent exposure, or maternal protection. We developed and analyzed one particular model for parasite densities that matches the features of malaria documented across multiple previous studies (Fig 5). The basic features of the function describe the time course of an infection: after an infectious bite, parasites spend around 6 days in the liver (18**?**); parasites enter the blood on or after day 7 of the infection and the asexual blood-stage parasites begin a period of geometric growth; symptoms can appear around day 9-10, and parasite populations become patent (*i*.*e*. detectable) around day 14; the geometric growth phase of an infection ends around day *δ* ≈ 20 days when parasite densities peak, and the infection begins a chronic phase during which parasite densities fluctuate overall but tend to decline as the infection ages. Infections can persist for several months, or even years; the duration of infections is highly variable. The model is described by the equation:

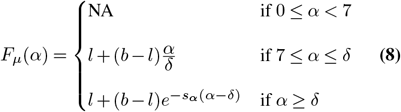

A key feature of the model is thus that infections have two distinct phases: an acute growth phase when parasite densities are increasing geometrically (for *α < δ*); and a chronic phase when parasite densities decline (Fig. 5). The parameter values here are provided as approximate values that are roughly consistent with observations, a starting point for further investigation about about which substantial uncertainty remains. This framework provides a rigorous way of formulating and testing ideas *in silico*.

### Clonal Infections

Total parasite densities in a clonal infection must be bounded between 1 and the total number of RBCs, which is a function of host size and age. The log_10_ number of RBCs is denoted *b*(*a*). We model Ω(*µ*), where *µ* is the expected value of log_10_ iRBCs (denoted *ξ*) as a function of the AoI; *µ* = *F*_*µ*_(*α*). To compute Ω, we modified a *beta* distribution to: 1) return a value between 0 and *b*(*a*); with mean log_10_ densities equal to *µ*; and 3) the variance, which computed as a function of the mean (Supporting Information, and `ramp.falciparum`). A random variable describing log_10_ parasite densities in each parasite clonal infection cohort is denoted *P*_*τ*_ (*a*), with a probability density function:

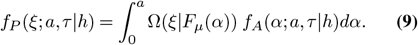

Fig 6. We let *F*_*P*_ denote the CDF, and 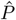 the random number function. The software, `ramp.falciparum`, was developed around a flexible design to accommodate alternative model formulations, (*i*.*e*., for both *F*_*µ*_ and Ω). Ω could thus be developed for any reasonable double bounded continuous probability distribution function family.

### Parasite Densities

To model parasite densities in complex infections, some assumption about interactions among parasites within a host is required. Do they compete for resources or immune-free space, interfere directly, facilitate one another or fluctuate independently? While there are many compelling candidate models that could be developed and tested, we have analyzed parasite densities under the simple and parsimonious assumption, consistent with the other assumptions of *M/M/*∞, that parasite densities fluctuate independently. Alternative models based on other assumptions can be explored in future studies. Computing total densities for potentially complex infections thus entails adding up the parasite densities of all parasite clones, the number of which is given by the distribution of the MoI.

Let *B* denote a random variable describing log_10_ parasite densities summed over all clonal infections in a host cohort of age *a*, denoted *B*_*τ*_ (*a*) with a probability density function:

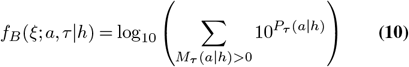

We let *F*_*B*_ denote the distribution function, and 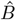 the random number function. We were unable to derive a simple formula for *B*_*τ*_ (*a*), but we have developed algorithms to compute it and to verify the computation (`ramp.falciparum`).

This equation can be understood through the pseudo-code that describes how to draw random values of parasite densities from individual hosts in a cohort of age *a* born on day *τ*, by drawing a random number of clonal infections with random values for clonal parasite densities (Algorithm 1).

[1] *m*_*τ*_ (*a*|*h*) Compute Mean MoI 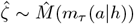 Draw Random MoI 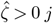 in 1 to 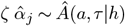 Draw Random 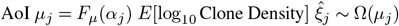Draw Random Clone 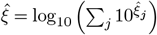 Sum of Clone Densities 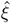 is Undefined Host is Uninfected

We developed functions to compute the density and distribution functions for complex infections, as a convolution over a distribution of MoI (Fig 7b, Fig 6c, `ramp.falciparum`, and Supporting Information). As an intermediate step, we developed random variables for parasite densities in infections with MoI= *N*, with distribution function *f*_*N*_ (*ξ*; *a, τ* | *h*), and functions to draw random number *N*-tuples from *P*_*τ*_ (*a*). We verified the convolution-based algorithms against these density and distribution functions in complex infections with fixed MoI (Fig 6c, Supporting Information, `ramp.falciparum`).

**Fig. 7.**
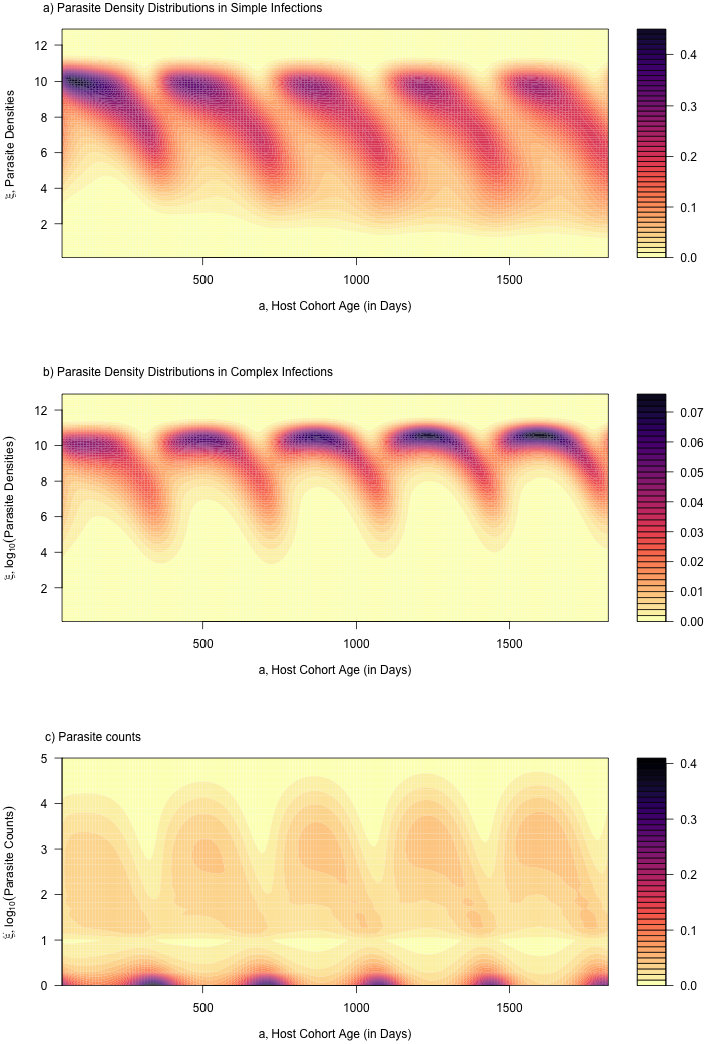
Parasite densities and counts for the same cohort illustrated in Fig. 4. a) The distribution of log_10_ parasite densities across all cohorts of parasites, log_10_ *P*; b) The distribution of log_10_ parasite densities in the host cohort as it ages, log_10_ *B*; and c) The distribution of log_10_ parasite counts, conditioned on counting at least one parasite, log_10_ *C*. Compare the patterns in panels a) and b) here to the patterns in panels b) and c) in Fig 4. Mean log_10_ parasite densities in complex infections can be understood in terms of the AoY.

Since *B* is a log_10_ transformed value of a sum of values of 10^*P*^, it follows that *B* is close to the maximum value of *P*, and we can bound the sum:

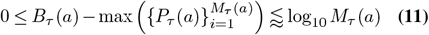

The total density would be near the upper bound only if all clonal parasite densities were approximately equal. If *N* clones had approximately the same density, then the total density would be *N* times the maximum value of P, but after log transforming parasite densities, the effect adds log_10_ *N*. For *N* = 2, parasite densities are approximately 0.3 higher in log_10_ parasite densities, and for *N* = 5, approximately 0.7 higher. Compared to the daily variance in parasite densities observed in malariotherapy patients (Fig 5), these numbers are small (8). Superinfection does have a big effect on parasite density distributions in this model, but mainly because parasite densities a sum of multiple infections, which is bounded below by the one with the highest density (See Fig 6c and compare Fig 7a and Fig 7b).

Since parasite density distributions fluctuate over several powers of ten, the sums of parasite densities can be guided by an understanding of extreme values. It follows that *B* is likely to follow the youngest infection, not the AoI, which motivated development of a new random variable describing the youngest infection (Fig 6).

A potential problem with this model formulation is that while the parasite densities for individual clones are bounded well below 10^*b*(*a*)^, total parasite densities summed over multiple clones need not be. To deal with extreme chance events in the upper tail, the mean of the distribution of parasite densities was bounded well below the total number of iRBCs, such that computed parasite densities exceeding 1% of all RBCs are extremely rare. Although this does break independence of infections, this distortion arises rarely and does not have a large impact on calculations on detection as this saturates quickly at high densities. Practically if parasite densities in a patient were close to the absolute maximum, catastrophic health risks would be likely. Hyperparasitemia, a form of severe malaria defined by having more that 5% infected RBCs in immunologically-naïve individuals, is associated with severe, acute clinical symptoms.

### Age of the Youngest Infection (AoY)

We can take advantage of the extreme value arithmetic that govern parasite density distributions, as suggested by Eq. 11, by developing a new random variable, *Y*_*τ*_ (*a*), that describes the age of the youngest infection (AoY) with the distribution function:

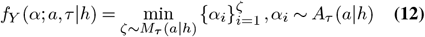

Logically, since the youngest infections tend to have the highest densities (unless a parasite is in the liver or early acute growth phase), expected parasite densities in infections would be dominated either by the *oldest* clone in the acute phase or the *youngest* parasite clone in the chronic phase. In this derivation of the AoY, we ignore the inaccuracies introduced by the early acute phase infections.

To compute *Y*, we derive a formula for the AoY in terms of the AoI density and distribution functions, and the MoI:

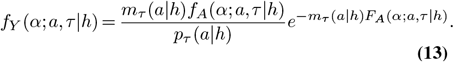

We let *F*_*Y*_ denote the CDF, and *Ŷ* the random number function.

Based on the arguments made above, we can use *Y* to compute a lower bound on *B*_*τ*_ (*a*), a distribution that would be observed if the entire distribution was determined by the youngest clone, 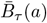 with a distribution function:

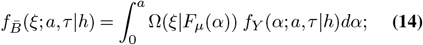

In effect, 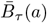 is computed by substituting *Y* for *A* (also see Fig. 6a,b), and

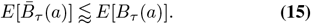

We note that the distribution of the AoY has a very different shape than the AoI (contrast Fig 4b,c, and see Fig. 5). While the approximation is not exact, it provides a useful heuristic; the shape of *Y*, not *A*, explains the shapes of the distribution of parasite densities for simple and complex infections (compare Fig 7b,c).

### Hybrid Variables

Using random variables, we can compute parasite density distributions in a cohort of any age given a function that describes the history of exposure (the FoI, *h*_*τ*_ (*a*)), an assumption about parasite clearance, *r*, and a function that describes parasite densities as infections age, Ω(*F*_*µ*_(*α*)). A disadvantage of the probabilistic approach is that it can be computationally intensive, and it is difficult integrate these equations into models for the transmission dynamics and control of malaria. To address these limitations, we developed new hybrid variables that are computationally much simpler (44). It has been shown that the dynamic changes in mean MoI, summarizing much of the behavior of the system of equations in Fig 1, can be modeled as a simple ordinary differential equation (44). Noting that *m*_*τ*_ (*a*) is the mean MoI, we also define the mean AoI, *x*_*τ*_ (*a*) and the mean AoY, *y*_*τ*_ (*a*):

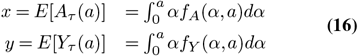

Next, we derived equations describing the dynamics of the variables *x* and *y* (Supporting Information), which when combined with the hybrid model for the MoI (44), allow us to compute changes over time using a simple system of differential equations:

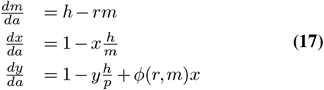

The function *ϕ*(*r, m*) captures the dynamics of changes in the AoY in complex infections, in which the loss of the youngest infection doesn’t result in clearance but an increase in the age of the youngest clone (see the Supporting Information).

Further, the mean MoI, AoI, and AoY can be used compute approximate mean logged parasite densities:

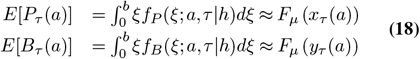

While the approximations are not exact, they are reasonably close (Fig. 8). These approximations, and the underlying heuristic, thus serves as a useful conceptual bridge connecting exposure to mean parasite densities.

**Fig. 8.**
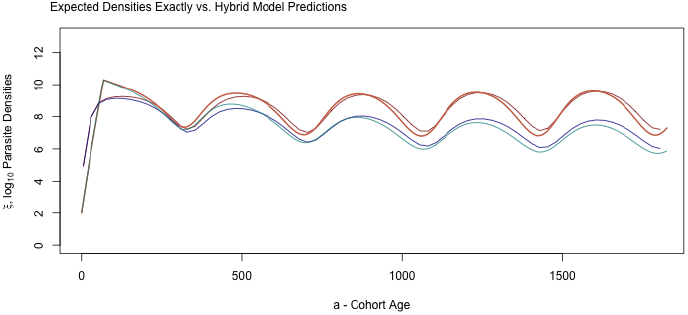
Mean log10 parasite densities (red hues) and clone densities (blue hues) computed exactly and using the hybrid variable approximations Eqs 18. In both cases, the hybrid variable approximation is the slightly lighter shade. The approximation performs comparatively poorly for the first 200 days because of the strongly non-linear relationship between the AoI and parasite densities around the peak. Thereafter, the approximations differ slightly from the exact values.

### Measuring Infection

The random variables describing parasite infections and parasite densities are unobservable (latent) states, so to understand malaria infections, we need to model the observation process. What is the relationship between our latent random variables and observable quantities? Studies in malaria epidemiology often rely on parasite prevalence or parasite counts. We thus developed random variables describing parasite counts from examination of a blood sample, from which we derived a related variable describing the probability of counting at least one parasite.

We developed random variables for parasite counts on light microscopy. Sources of sampling error include variation in parasite densities in the sampled blood, and errors introduced through counting, which involves examination of a set of fields on the blood slide (48). These differences can be affected by differences among light microscopists. We note, as an aside, that we have not considered a set of issues associated with false positives.

Similar kinds of concerns would apply to modeling PCR or RDTs, but new random variables would need to be developed around a sampling model that was adapted to the diagnostic method. It is reasonable to expect that the fraction of samples that were positive would be different based on the sampling model as well as the sensitivity of the method (*e*.*g*., by light microscopy, qPCR or rapid diagnostic tests which detect a biomarker that is believed to be strongly correlated with parasite densities).

### Sampling

To diagnose malaria by light microscopy, a small fraction of total blood is sampled and examined for parasites. Parasites are thus often not detected, even when they are present. The relationship between the true parasite densities and parasite counts depends on the total fraction of blood examined, *q*. For light microscopy, where approximately 1 ≈ *µ*l, of blood is drawn and a fraction of it examined. If *s* ≈ 6 were the log number of RBCs in a blood sample, then the fraction of RBCs examined would be approximately:

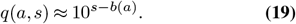

The log_10_ parasite counts in a sample, 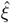,is a random variable that depends on the log_10_ parasite densities in blood, *ξ*. We let *S* denote a probability density function family to model raw counts as a function of parasite densities, conditioned on being infected. We let *Q* denote the probability of counting at least one parasite, conditioned on sampling an infected host:

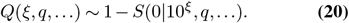

Next, we define a function describing the distribution of log_10_ positive counts (*i*.*e*., conditioned on detecting at least one parasite)

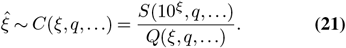

In developing models for counts in complex infections, we identified some issues that we could not resolve from *a priori* considerations: What probability density function family is appropriate for sampling parasites and parasite counts data? and should counts be modeled for independent detection of individual parasite clones, or for aggregate parasite densities in an infection?

On the choice of sampling distributions, if blood were drawn and parasites counted repeatedly, would the counts data follow a Poisson distribution, negative binomial distribution, or some other probability distribution function family? On the question of whether we should treat parasite densities in the aggregate, or whether to treat clones independently, we could find no strong *a priori* reason to prefer one or the other, so we developed both methods, which very similar, but slightly different predictions. In the associated software, `ramp.falciparum,` functions were developed to model detection based on a Poisson or negative binomial family of distributions, and the software design is extensible, making it possible to add other suitable probability density function families. In the following, we use the negative binomial distribution.

We fit a model for parasite density distributions and detection to the malaria-therapy data using maximum likelihood estimation (Supplement-S5, Fig 9). The analysis gave different answers depending on the value of the size parameter of the negative binomial parameter, and parameters describing the variance in parasite population densities among individuals as a function of the mean. If the size parameter was larger than one, then the sampling was reasonably accurate, andwparasite densities among individuals were more variable. If the size parameter was smaller (≈ 0.2), then the sampling was extremely noisy, and parasite population densities were less variable. A few studies that resampled infected individuals at short intervals suggested the sampling is reasonably accurate (22, 23).

**Fig. 9.**
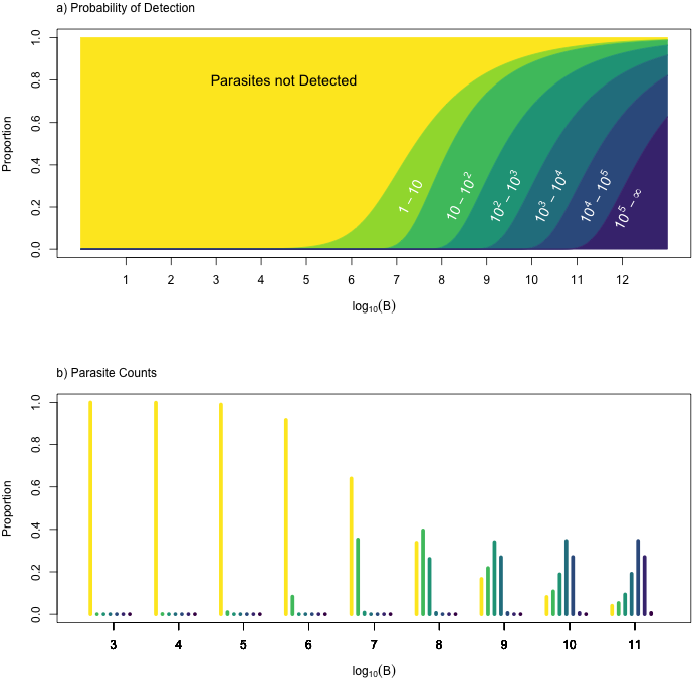
Sampling error for the detection process. a) For values of log_10_ parasite densities (*x*-axis), vertical slices show the proportion of parasite counts in a range defined by powers of 10 (*y*-axis). To make this plot, we assume the proportion of blood examined is *q* = 10^−6^, and the size parameter was fitted to the malariatherapy data. b) For each integer value of *B*, we show a histogram of a vertical slice of the figure above it. Each one is a histogram, binned by powers of 10 (colors and categories match the top figure) for log_10_(*B*) for integer values from 3 to 11.

**Fig. 10.**
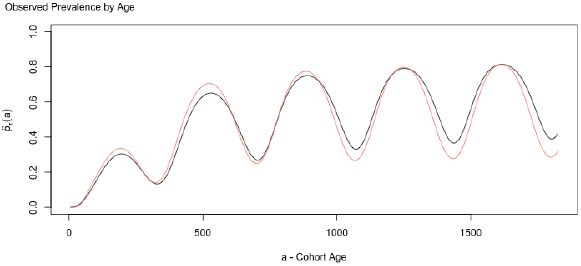
The prevalence over time computed using random variables (Eq. 23), and an approximation using hybrid variables (Eq. 28). The errors in this approximation might be too large to be useful, but the comparison is a useful heuristic for understanding detection, and it can serve as a basis for translating hybrid variables into observations using the probabilistic framework.

**Counts**

For detection based on aggregate parasite densities, we can simply use *C* to compute parasite counts over the distribution of parasite densities, *B*_*τ*_ (*a*) (*e*.*g*. see Fig 9):

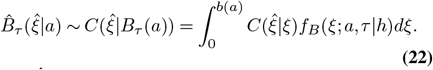

Since 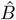 *B* describes the distribution of zero counts, and positive counts for slides from infected hosts, the number of zero counts over all slides examined is thus inflated by *p*_*τ*_ (*a*).

### Detection

Using *B*_*τ*_ (*a*), apparent prevalence is

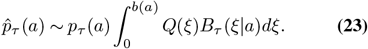

Alternatively, we developed a model around detection of independent clones, which aligns in spirit with the model *M/M/*∞. We define a function, *F*_*Q*_, that computes the probability of detection as a function of the age of infection *α*, using the density distribution function, Ω and *F*_*µ*_:

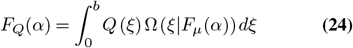

If present, a clone would be detected with probability:

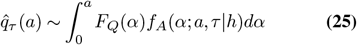

so the apparent MoI, 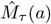,if individual clones could be counted, would be Poisson distributed with mean 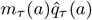 and alternative formula for detection of parasites in a complex infection is thus:

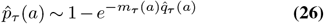

### Hybrid Detection Dynamics

We can also show that mean logged parasite counts for parasite clones scale in a reasonably simple way,

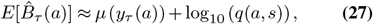

with interesting counts distributions (Fig 7c). We can also show that we can compute a good approximation to the observed prevalence using hybrid variables:

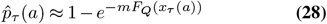

Once again, these approximations are useful as a heuristic to explore understanding the relationship between true and observed prevalence.

## Discussion

Using a probabilistic framework, we showed how exposure over time translates into patterns of infection and malaria parasite densities and prevalence in cohort of children as they age. The approach emphasizes that malaria infection and all the metrics we use to measure it hold a memory of recent exposure. Since parasite densities are highly stochastic, the memory of exposure is probabilistic, observable only by sampling from cohorts in a population. We also showed that this memory is well-approximated by a simple system with hybrid variables that describe the mean MoI, AoI and AoY. The quantitative logic of these two approaches, relating probabilistic models to hybrid variables, makes it possible to draw a clear line from exposure to parasite density distributions. In the statistical sense, parasite densities are well-predicted by the AoY. While the quantitative logic connecting the two is long and sometimes mathematically cumbersome, it illustrates how a detailed and highly realistic probabilistic description of a system with variables captures much of the complexity. While the models we have presented lack some of the core features of malaria – immunity, disease, treatment and chemoprotection, and infectiousness – these challenges can also be handled by extending the probabilistic approach. The approaches are planned for future studies. Altogether, this synthesis represents a new way of understanding malaria epidemiology represents a new way of understanding malaria epidemiology.

Although the medical approach to understanding malaria epidemiology relies heavily on parasite densities, it is difficult to understand patterns without grappling with the large uncertainty associated with this data and the underlying process. Without doing so, we are likely to overlook existing but messy relationships such as that between malaria incidence and febrile infections in the growth phase. In developing the functions, *µ*(*α*), our attention was drawn to a minor, but perhaps critically important feature of infections: some features of malaria epidemiology could occur early in the growth phase, while parasite densities are still low (8). There is a narrow window of time, probably less a week, in which the growth phase of a parasite was observed to cause disease, but when parasite densities are comparatively low (8). Severe disease may be therefore be associated with these growth-phase acute infections. The observation begs a serious question about using parasite densities as a covariate in studies of malaria. Symptoms that arise from parasite infections in the acute phase may not undetectable or they present with low densities, while parasite densities from old infections may be a source of noise.

This study takes a semi-mechanistic approach that starts from the premise that parasite densities can be computed, in a statistical sense, using information about infection age. The probabilistic nature of the model acknowledges the nature of infection age as predicting - not determining - the parasitemia in an individual chosen at random. Because we wished to model observable quantities such as measured prevalence 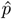 which depend on the latent state of the parasitemia in an infected individual chosen at random, it was not enough to know the expectation alone. As we model each individual as being equally likely to be measured here for simplicity, the predicted rate of detection - a function of parasitemia - must be weighted according to its likelihood of being observed. In order to model variables which can be predicted through other functions of parasitemia, such as the probability of fever, more information about the parasite density distribution may be required. The approach does not address the question of what drives the shape of curves describing parasite densities as they age, but by finding functions that emulate the behavior of other models, including individual-based within-host models, it could provide a rigorous basis for comparison.

While there are many parameters in the full model, each sub-model is parametrically simple. To describe parasite densities with a high degree of realism, we have we developed several linked sub-models, each one of which is highly realistic. The dynamics of infection under the model *M/M/*∞ have one fixed parameter (*r*) describing the loss of infections, a decay rate that describes how prevalence reflects the *memory* of past exposure. The model for parasite densities and detection requires the most: five parameters describing the trajectory of the expected value (two parameters are bounds, one describes time in the liver, one describes parasite clonal density at peak, and one describes the decline in parasite densities with the parasite clone’s age. The distribution function required three parameters to express the variance as a function of the mean. Two parameters were needed to describing detection, of which one is context dependent (*i*.*e*. fraction of blood examined). The methods and the software can be extended to other mathematical forms.

In presenting this framework, we emphasize that the claims are not specific inferential claims but general ones. The general claim is that models of this sort naturally replicate the sorts of quantitative patterns that have been observed in observational malaria epidemiological studies. The claims are not about any specific mechanism, but the method provides a useful way of understanding and modeling malaria epidemiology. Since the semi-Markov models are extending an observed pattern, it is not surprising that they should give seemingly reasonable results.

The approach taken here also emphasizes an a posteriori approach with a priori tools (49). That is, after observing patterns in the data for parasitemia which is difficult to model (a posteriori), we created an a priori model of infection age and used the statistical model linking it to parasitemia to then produce a simpler model of parasitemia. To emphasize the structure and flow of the model, we largely implemented computational approaches here. Approximations suggest some robustness in the structure of the models as output is weakened only slightly by using the age of the youngest infection.

Many of the complex features of malaria epidemiology are made more coherent when seen as emergent properties of the biological needs of the malaria parasite. Sexual reproduction occurs in the mosquito, and outcrossing occurs only if the multiplicity of infection (MoI) is greater than one. Frequent outcrossing, in turn, supports a genetically diverse parasite population, which is partly responsible for the weakness of the immune responses and other features of malaria epidemiology.

Most outcomes of being exposed to malaria are expected to vary with an individual’s age and previous history of exposure and infection. The core challenges for applied malaria epidemiology are to understand or predict likely outcomes and to identify modifiable factors affecting the burden of malaria and the prospects for malaria elimination. Framed broadly, outcomes are affected by host infection status (*e*.*g*. parasite densities), acquired immunity resulting from a lifetime of exposure, other host factors (anemia, genetics, nutrition, *etc*.), parasite factors (genotype and phenotype), and case management with anti-malarial drugs. The difficulty of understanding and quantifying outcomes is increased by the challenge of simultaneously measuring recent exposure, infection, disease, immunity acquired over a lifetime of exposure (*e*.*g*. through serology), and other important covariates in enough settings to quantify their relationships. The contemporary view of malaria epidemiology has been pieced together from a heterogeneous set of studies conducted over roughly 140 years, but the number of studies that has been done is small compared to the number that would be needed. Important gaps in our understanding of malaria epidemiology remain.

## Conclusion

Parasite infection dynamics and detection are but a starting point for understanding malaria epidemiology. We have introduced a new modeling framework which bridges the gap between stochastic individual-level behavior and population-level observable quantities. This is a novel simulation method for infectious disease that takes a non-compartmental approach to managing the complexity and uncertainty inherent in the mechanisms that control parasitemia. However as with all models, this complexity and stochasticity can hinder our ability to isolate mechanism almost as much as the system it is meant to model. Future directions include extending the framework to model immunity, disease, treatment, and transmission. We also plan to explore the effects of disease-prompted treatment on age-prevalence curves; and comparing the evolutionary dynamics of the parasite with strain-specific immunity.

## Data Availability

All data and code are available online from:
https://dd-harp.github.io/ramp.falciparum/
or
https://github.com/dd-harp/memory.core

https://github.com/dd-harp/memory.core

## Author Contributions

JMH and DLS designed the study, carried out all the analyses. JMH, DLS, ARC, and SLW wrote the first draft. All authors contributed to the final draft.

## ACKNOWLEDGEMENTS

This research was supported by a grant from the National Institute of Allergies and Infectious Diseases (R01 AI163398), which supported DLS, JMH, and ARC. Additional support for DLS from the Bill and Melinda Gates Foundation (INV 030600).

